# Thalamic tFUS for Post-Stroke Motor Recovery: A Pilot Multimodal Neurobehavioral Study

**DOI:** 10.64898/2026.07.07.26357338

**Authors:** Simeng Wu, Xin Zhang, Jixuan Kang, Yan Chen, Heng Wang, Hehe Chen, Luhan Zhang, Weixin Zhu, Xiao Zhang

## Abstract

Effective modulation of cortical–subcortical motor circuits is essential for post-stroke recovery, yet progress has been constrained by the absence of non-invasive tools capable of precisely targeting deep brain structures. In this pilot proof-of-concept study, we explored the feasibility and preliminary neuromodulatory effects of a 12-minute transcranial focused ultrasound (tFUS) protocol targeting the ipsilesional ventral lateral posterior (VLp) thalamus in ischemic stroke patients. Six individuals with upper-limb hemiparesis received individualized, neuronavigation-guided tFUS. Sensorimotor tracking performance improved significantly after a single session. Concurrent EEG revealed reversible beta-power suppression over the ipsilesional motor cortex and enhanced theta-phase synchronization in frontoparietal networks, both of which were associated with behavioral gains. Resting-state fMRI indicated rebalancing of inter-hemispheric motor networks. These preliminary findings suggest that thalamic tFUS can modulate both local and network-level neural activity and is associated with immediate functional improvement, highlighting its potential as a feasible neuromodulation approach for deep motor circuit engagement in post-stroke rehabilitation.

## I. Introduction

Stroke remains one of the leading causes of long-term disability worldwide [1], with motor impairments affecting a significant proportion of survivors. Functional recovery relies on brain neuroplasticity [2], which involves the reorganization of cortical and subcortical networks [3]. A major obstacle, however, lies in the limitations of current neuromodulation tools [4]. Non-invasive techniques such as transcranial magnetic stimulation (TMS) and transcranial direct current stimulation (tDCS) have demonstrated variable therapeutic efficacy [5], [6], largely due to their limited spatial resolution and inability to reliably access deep motor control structures [7].

Transcranial focused ultrasound (tFUS) has recently emerged as a promising technology to overcome this barrier [8]. It delivers focused acoustic energy with high spatial precision, enabling non-invasive, millimeter-scale neuromodulation of deep subcortical structures without substantially affecting the overlying cortex [9]. Despite growing interest, the application of tFUS in stroke populations remains limited. Existing studies have primarily targeted cortical regions, particularly the primary motor cortex (M1) [10], [11]. A compelling therapeutic target within the motor network is the thalamus—specifically the ventral lateral posterior (VLp) nucleus, which serves as a critical relay hub [12] within the cortico–basal ganglia–thalamocortical loop [13]. Following stroke, disrupted thalamocortical communication is implicated in impaired motor function and maladaptive inter-hemispheric inhibition [14]. Although tFUS has yielded promising results in modulating cortical and deep brain regions in both animal models and early human studies [15], [16], the underlying neural mechanisms through which such intervention exerts its effects—particularly in the disrupted post-stroke network— remain poorly understood.

To address these gaps, we conducted a pilot multimodal study to explore the feasibility and preliminary neurobehavioral responses to precisely targeted thalamic tFUS in patients with chronic stroke. Specifically, we assessed (1) the immediate effect on sensorimotor integration, (2) the underlying online neural mechanisms via concurrent EEG, and (3) the subsequent reorganization of large-scale motor networks using resting-state fMRI. This pilot study was designed as a necessary first step to evaluate the technical feasibility of VLp-targeted tFUS in stroke and to gather initial mechanistic insights that could inform the design of future controlled trials.

## II. Materials and Methods

### A. Participants

We enrolled six male patients (age 57 ± 11.15 years) with a first-ever ischemic stroke and unilateral upper-limb motor impairment. Stroke onset ranged from 52 to 103 days (Table I). Exclusion criteria included MRI contraindications, thalamic lesions, severe cognitive impairment, other neurological conditions affecting motor function, and medications influencing cortical excitability. Written informed consent was obtained from all participants.

**TABLE I.** Demographic Information of Participants.

| ID | Age | Hemisphere* | Days after stroke | Barthel scale |
| --- | --- | --- | --- | --- |
| 1 | 51-55 | Left | 101-125 | 70 |
| 2 | 61-65 | Right | 50-75 | 55 |
| 3 | 61-65 | Left | 76-100 | 70 |
| 4 | 41-45 | Right | 50-75 | 55 |
| 5 | 51-55 | Left | 76-100 | 75 |
| 6 | 71-75 | Left | 50-75 | 35 |

\*Hemisphere refers to the lesional brain hemisphere.

### B. Overall Study Design

The experimental protocol consisted of three phases: a comprehensive baseline assessment phase and two tFUS intervention phases, enabling the evaluation of online and short-term effects, respectively (Fig. 1A). During the baseline session, structural CT/MRI scans were acquired for individualized target segmentation and acoustic simulation. A 10-minute resting-state fMRI scan and the pursuit-tracking paradigm [17] were administered to establish baseline measurements. In the pursuit-tracking task, participants moved a mouse to align the cursor with a randomly moving target; the vertical tracking error served as the primary outcome metric.

**Fig. 1.**
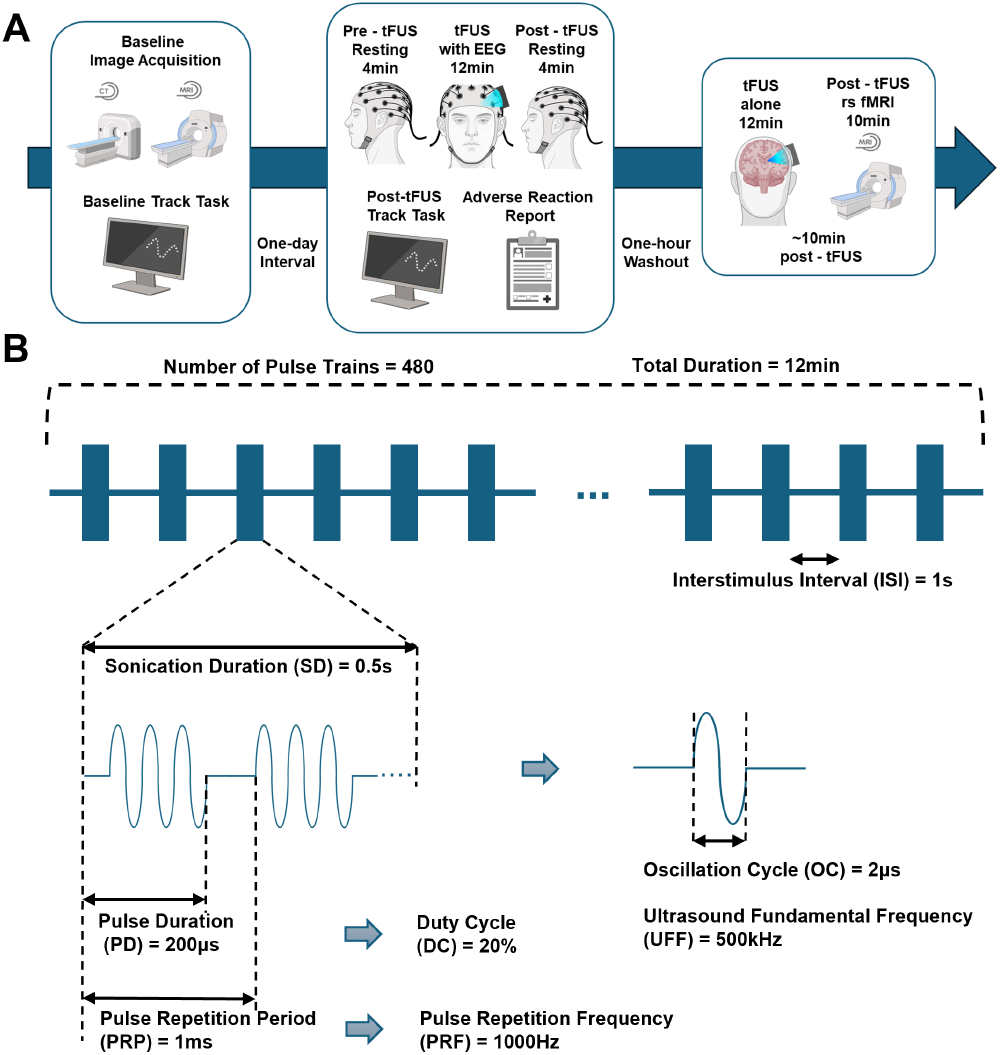
Procedure and Protocol. (A) Experimental procedure and timeline. (B) Parameters of the tFUS protocol.

Following a one-day interval, a 12-minute tFUS session was delivered with concurrent EEG to capture online neural activity (including 4-min resting-state EEG pre- and post-tFUS). The post-tFUS pursuit-tracking task was administered immediately afterward to assess behavioral changes, and participants were interviewed regarding adverse effects. Subsequently, a second 12-minute tFUS session (tFUS-only) was conducted, immediately followed by a 10-minute resting-state fMRI scan. The two tFUS sessions were separated by a minimum one-hour washout period.

### C. tFUS Protocol

We used the NeuroFUS TPO and DPX-500 transducer (Brainbox Ltd., Cardiff, UK), a four-element ultrasound transducer with a central frequency of 500 kHz. Stimulation parameters were set to a pulse repetition frequency (PRF) of 1000 Hz, a duty cycle (DC) of 20%, a sonication duration of 0.5 s, and an inter-stimulus interval (ISI) of 1 s. Throughout the 12-minute stimulation period, a total of 480 pulse trains were delivered (Fig. 1B). The in situ spatial-peak pulse-average intensity (I_SPPA_) was maintained at 8 W/cm^2^ for all participants. To accommodate the transducer, a customized EEG cap was used. The participant’s head and the transducer assembly were stabilized using an integrated system of adjustable straps, a mechanical arm mount, and a chin rest.

### D. EEG Acquisition and Preprocessing

EEG was recorded using the 64-channel SynAmps RT system (Neuroscan) at a sampling rate of 1000 Hz. Electrode impedances were maintained below 10 kΩ with conductive electrolyte gel. Data were downsampled to 500 Hz and bandpass filtered (1–80 Hz) to attenuate low-frequency drift and high-frequency noise, followed by a 50 Hz notch filter for line noise removal. Channels exhibiting poor signal quality were rejected and interpolated using spherical spline interpolation. Artifacts arising from ocular, muscular, and other non-neural sources were removed via independent component analysis (ICA). All data were visually inspected to verify artifact removal.

### E. MRI Acquisition and Preprocessing

MRI data were acquired on a 3T Philips Achieva scanner with an 8-channel SENSE head coil. High-resolution structural images were obtained using T1-weighted 3D turbo field echo (TFE) and T2-weighted 3D turbo spin echo (TSE) sequences for co-registration and normalization. Functional images were acquired with a T2*-weighted echo-planar imaging (EPI) sequence. Key acquisition parameters included: T1w TFE (TR = 600 ms, TE = 28.3 ms, voxel size = 1× 1× 1 mm^3^), T2w TSE (TR = 3000 ms, TE = 243.1 ms, voxel size = 1 ×1× 1 mm^3^), and resting-state EPI (TR = 2972.64 ms, TE = 35 ms, voxel size = 3× 3 ×3 mm^3^). Preprocessing included (a) removal of the first five volumes, (b) realignment, (c) slice-timing correction, (d) artifact detection (framewise displacement > 0.9 mm or global BOLD signal > 5 SD), (e) segmentation, and (f) normalization to MNI space using the IXI-549 template. Images were resampled to 2 mm isotropic voxels and smoothed with an 8 mm FWHM Gaussian kernel.

### F. Statistical Analysis

Paired *t*-tests were applied to compare baseline and post-tFUS behavioral and fMRI data, as well as to compare the pre-, during-, and post-tFUS EEG data, given the within-subject design of the study. For EEG, spectral power and phase-locking value (PLV) were calculated separately for each frequency band, with false discovery rate (FDR) correction applied across all EEG channels and frequency bands to control for multiple comparisons. For fMRI data, voxel-wise analysis was conducted using a general linear model (GLM) in SPM12, with initial uncorrected thresholds at *p* < 0.001, followed by cluster-level family-wise error (FWE) correction at *p* < 0.05. Statistical significance for all EEG and fMRI comparisons was set at *p* < 0.05 after correction. Given the small sample size (*N* = 6), all correlation analyses should be interpreted with caution, as they are susceptible to the influence of individual data points.

## III. Results

We assessed the feasibility of a novel 12-minute tFUS protocol targeting the VLp thalamic nucleus for post-stroke hemiparesis. To ensure spatial consistency across participants, the left hemisphere was operationally defined as the lesional hemisphere for all analyses; consequently, EEG channels and fMRI images were mirrored left–right for participants with right-hemisphere lesions. We examined the underlying neurobehavioral changes, integrating measures of motor performance, neural oscillatory dynamics, and large-scale network reorganization.

### A. Effective Targeting of the VLp Nucleus with tFUS

To ensure precise targeting, the VLp nucleus was individually localized and segmented for each participant using baseline CT and high-resolution T1- and T2-weighted MRI. The BabelBrain software was subsequently used for tFUS beamforming and acoustic field simulation (Fig. 2). Acoustic field simulations were conducted to model the distribution of ultrasound intensity and the resultant thermal effects. The intensity simulation estimated an in situ intensity of 8 W/cm^2^ at the target, while the thermal simulation predicted a maximum temperature rise of 0.2 °C, confirming the safety of the acoustic exposure. No adverse reactions were reported.

**Fig. 2.**
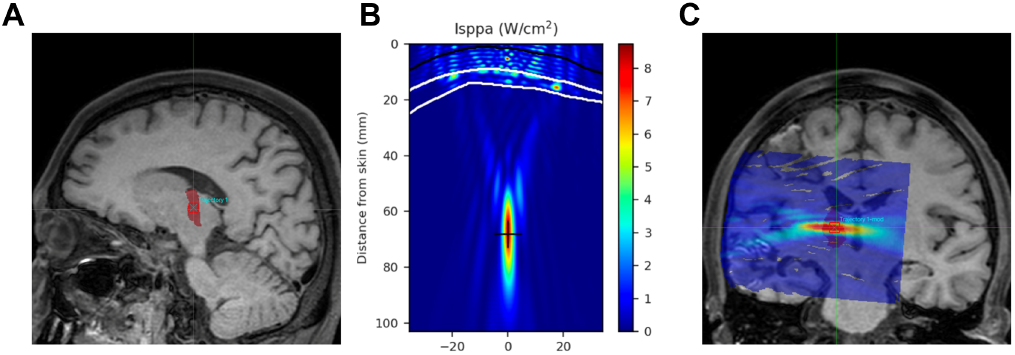
Target Segmentation and Acoustic Simulation. (A) Segmentation and positioning of the VLp, demarcated by the red shaded region on the individual’s anatomical scan. (B) Intracranial sound field simulation, ensuring that the target is located at the focal center of the sound beam. (C) Intracranial acoustic field mapped onto the individual’s anatomical scan. The VLp target is colocalized with the center of the tFUS beam.

### B. Behavioral Improvement in the Pursuit-Tracking Task

To assess the functional effect of tFUS, pursuit-tracking task performance after tFUS was compared with the baseline. Representative trajectory recordings from a baseline trial and a post-tFUS trial are displayed in Fig. 3A and 3B, respectively, with the target path shown in red and the recorded cursor trajectory in blue. Greater overlap between the curves reflects improved tracking accuracy after tFUS. A paired-sample *t*-test confirmed a significant reduction in vertical error following tFUS compared to baseline (Fig. 3C; *t*(5) = −4.54, *p* = 0.006, Cohen’s *d* = 1.85), suggesting an immediate improvement in sensorimotor integration. These preliminary results indicate a consistent positive trend in behavioral performance following thalamic tFUS.

**Fig. 3.**
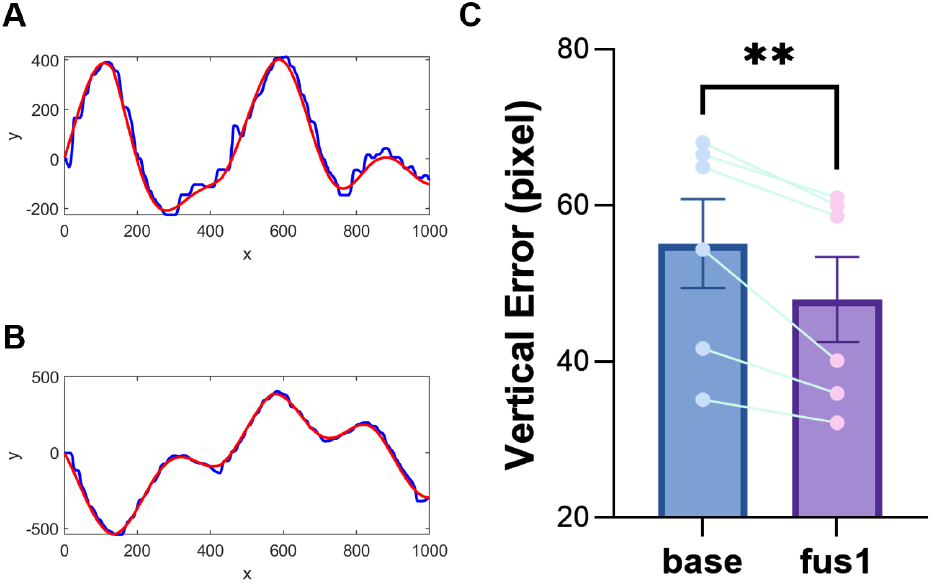
Pursuit-Tracking Task Performance. Representative trajectory recordings from a baseline trial (A) and a post-tFUS trial (B). Target path is shown in red and the cursor trajectory in blue. (C) Vertical error between baseline and post-tFUS Pursuit-Tracking task (^**^*p* < 0.01), Cohen’s *d* = 1.85.

### C. tFUS-Induced Online Neural Responses

Targeted VLp thalamic tFUS modulated local cortical activity and network-level synchronization. The key electrophysiological findings are summarized in Table II.

**TABLE II.** Summary of eeg Measures Across Experimental Epochs.

| Measure & Contrast | $t(5)$ | $p_{FDR}$ | Cohen’s $d$ |
| --- | --- | --- | --- |
| *Beta-band power at CP3* |  |  |  |
| fus – post | 11.54 | 0.004 | 4.71 |
| *Theta-band PLV (P3–FCz)* |  |  |  |
| fus – pre | 6.72 | 0.033 | 2.74 |
| fus – post | 12.15 | 0.032 | 4.96 |
| *Theta-band PLV (P2–F1)* |  |  |  |
| fus – pre | 11.74 | 0.023 | 4.79 |
| fus – post | 23.12 | 0.004 | 9.44 |

Absolute beta-band power at electrode CP3 (over the ipsilesional motor cortex) was significantly reduced during tFUS compared to the post-tFUS epoch (Fig. 4A–4B), indicating a reversible suppression. Beta oscillations (13–30 Hz) are associated with sensorimotor processing and motor inhibition; their reduction during stimulation suggests cortical disinhibition, which may facilitate motor preparation. A positive correlation was observed between the tFUS-induced reduction in beta power at CP3 and the behavioral improvement in the pursuit-tracking task (Fig. 4C; *r* = 0.871, *p* = 0.024). While this association is suggestive, it should be interpreted cautiously given the small sample size (*N* = 6), as individual data points can disproportionately influence the correlation estimate.

**Fig. 4.**
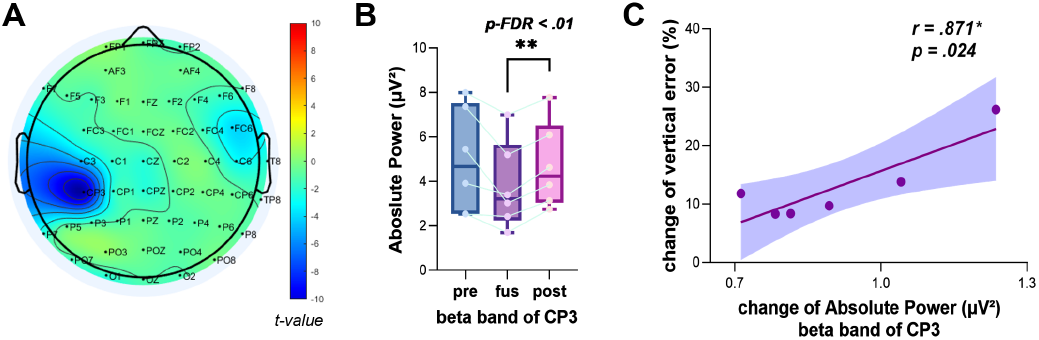
(A) T-map for the difference of absolute power (tFUS*−* post) in the beta band across all channels. (B) Absolute power in the beta band at channel CP3 during pre-, during-, and post-tFUS epochs (^**^*p*_FDR_ < 0.01). (C) Correlation between the change in absolute beta-band power at CP3 from tFUS to post-tFUS and the change in vertical error (*r* = 0.871, *p* = 0.024).

Simultaneously, tFUS enhanced network-level theta-phase synchronization between frontoparietal electrodes (Fig. 5). This theta-band PLV enhancement was also positively associated with behavioral gain (Fig. 5C; *r* = 0.874, *p* = 0.023), though the same caveat regarding sample size applies.

**Fig. 5.**
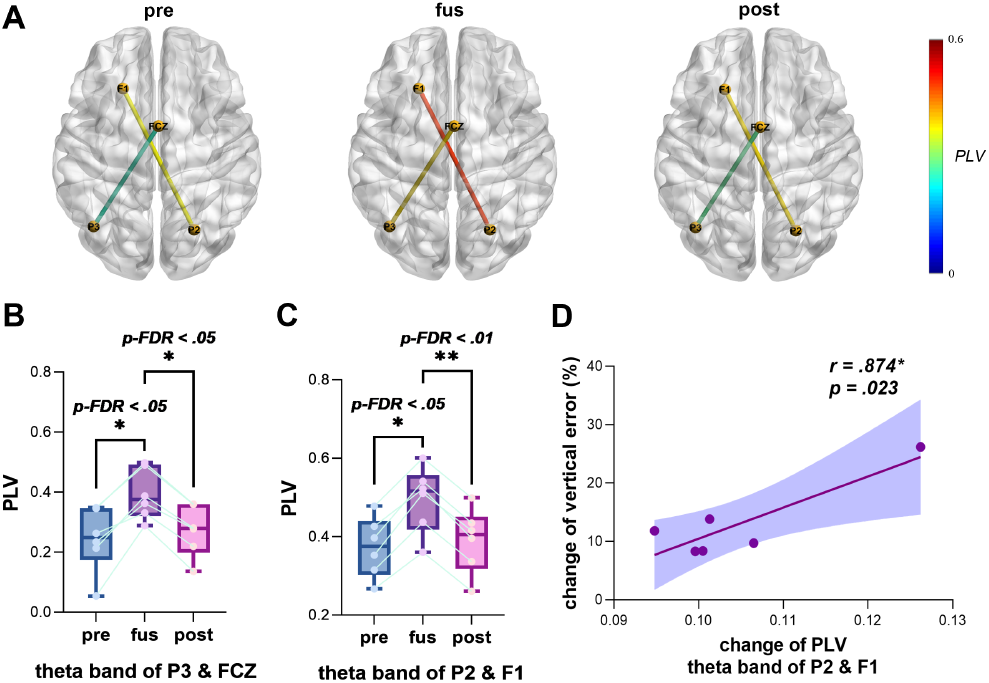
(A) PLV in the theta band during pre-tFUS (left), tFUS (middle), and post-tFUS (right) epochs. (B) PLV in the theta band at P3–FCz (^*\**^*p*_FDR_ < 0.05). (C) PLV in the theta band at P2–F1 (^*\**^*p*_FDR_ < 0.05, ^**^*p*_FDR_ < 0.01). Correlation between the change in PLV in the theta band at P2–F1 from tFUS to post-tFUS and the change in vertical error (*r* = 0.874, *p* = 0.023).

### D. tFUS Rebalances Inter-Hemispheric Motor Networks

Resting-state fMRI analysis revealed a baseline pattern of inter-hemispheric pathological imbalance, characterized by negative functional coupling between the bilateral thalamus and ipsilesional motor cortex (Fig. 6A). After tFUS, both regional homogeneity (ReHo) and amplitude of low-frequency fluctuations (ALFF) showed reduced hyperactivity in the contralesional primary motor cortex and supplementary motor cortex (Fig. 6B). In parallel, seed-based connectivity analysis demonstrated strengthened coupling between the bilateral sen-sorimotor network and the ipsilesional primary somatosensory cortex (Fig. 6B). These changes collectively suggest that tFUS facilitates the renormalization of motor networks from a pathological compensatory pattern toward a more ipsilesional-dominated reorganization.

**Fig. 6.**
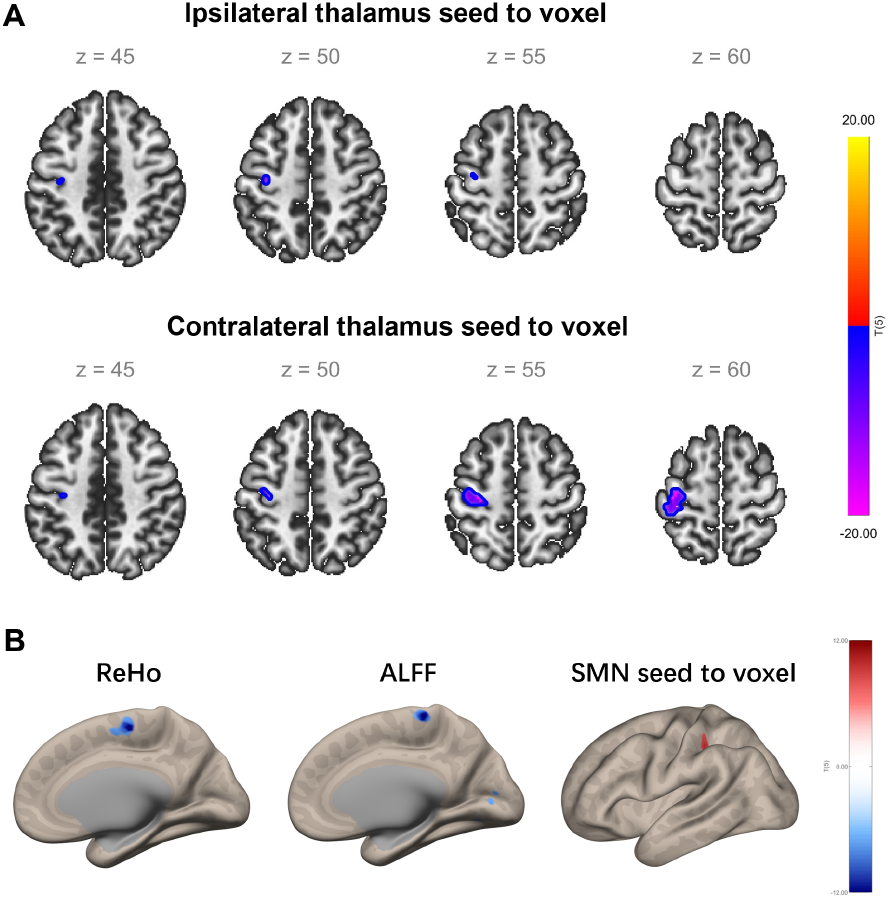
Functional connectivity at baseline and changes after tFUS (seed-based connectivity analysis), Regional Homogeneity (ReHo), and Amplitude of Low-Frequency Fluctuations (ALFF). (A) Whole-brain maps illustrating regions showing negative functional connectivity with the ipsilateral and contralateral thalamus seed applied to the ipsilesional motor cortex (24 voxels, center at *−*34, *−*16, +50; 165 voxels, center at *−*34, *−*20, +58). (B) ReHo decreased in the contralesional supplementary motor cortex (35 voxels, center at +6, *−*10, +60) and primary motor cortex (4 voxels, center at +6, *−*14, +60). ALFF decreased in the contralesional primary motor cortex (24 voxels, center at +4, *−*28, +66). Whole-brain maps illustrating regions showing increased functional connectivity with the bilateral sensorimotor network (SMN) seed after tFUS applied to the ipsilesional primary somatosensory cortex (23 voxels, center at *−*38, *−*32, +42), compared with baseline.

## IV. Conclusion

This pilot study demonstrates the initial feasibility and provides preliminary evidence for the neuromodulatory effects of tFUS targeting the VLp thalamic nucleus in stroke patients. Our findings reveal a pattern of local thalam-ocortical disinhibition—reflected in reversible beta-power suppression—accompanied by enhanced large-scale network communication, as evidenced by increased theta-phase coupling and inter-hemispheric rebalancing following stimulation. This multilevel modulation aligns with emerging models of tFUS action that span membrane-level effects to system-wide connectivity changes. Notably, while prior human tFUS research has largely focused on cortical targets, the present work extends the therapeutic focus to a deep motor relay hub and provides concurrent electrophysiological and neuroimaging evidence linking thalamic stimulation to functionally relevant neural changes.

Several critical limitations must be acknowledged. First, the small sample size (*N* = 6) limits statistical power and the generalizability of the findings; in particular, the reported brain–behavior correlations may be susceptible to undue influence from individual data points and should be regarded as exploratory. Second, the single-arm design without a sham or placebo control precludes definitive attribution of the observed effects to tFUS, as spontaneous recovery, practice effects, and placebo responses cannot be ruled out. These limitations are inherent to the proof-of-concept nature of this investigation and underscore the need for cautious interpretation. Nonetheless, this work establishes the technical feasibility of individualized VLp-targeted tFUS in stroke patients, offers a testable experimental framework, and provides the methodological foundation for future rigorous, sham-controlled trials with larger cohorts aimed at evaluating therapeutic efficacy, with the potential to advance deep neuromodulation paradigms in post-stroke rehabilitation.

## Data Availability

All data produced in the present study are available upon reasonable request to the authors.

## Acknowledgments

This work was supported by Noncommunicable Chronic Diseases-National Science and Technology Major Project (2024ZD0524402), Heilongjiang Provincial Natural Science Foundation Key Project (ZD2025H006). The experimental procedures involving human subjects described in this paper were approved by the Medical Ethics Review Committee of Jinhua Central Hospital (approval no.: 2024-208). The study was registered (ChiCTR2600121992).

## Notes

### Competing Interest Statement

The authors have declared no competing interest.

### Clinical Trial

ChiCTR2600121992

### Author Declarations

Medical Ethics Review Committee of Jinhua Central Hospital of Jinhua Central Hospital gave ethical approval for this work

